# Viruses previously identified in Brazil as belonging to HIV-1 circulating recombinant form (CRF) 72_BF1 represent two closely related CRFs, one of which, designated CRF122_BF1, is also circulating in Spain

**DOI:** 10.1101/2022.02.09.22270665

**Authors:** Javier E. Cañada-García, Elena Delgado, Horacio Gil, Sonia Benito, Mónica Sánchez, Antonio Ocampo, Jorge Julio Cabrera, Celia Miralles, Elena García-Bodas, Ana Mariño, Patricia Ordóñez, María José Gude, Carmen Ezpeleta, Michael M Thomson

## Abstract

Circulating recombinant forms (CRFs) are important components of the HIV-1 pandemic. Those derived from recombination between subtype B and subsubtype F1, with 18 reported, most of them of South American origin, are among the most diverse. Here we identify a HIV-1 BF1 recombinant cluster that is expanding in Spain, transmitted mainly via heterosexual contact, which, analyzed in near full-length genomes in 4 viruses, exhibited a coincident BF1 mosaic structure, with 12 breakpoints, that fully coincided with that of two viruses from Brazil (10BR_MG003 and 10BR_MG005), previously classified as CRF72_BF1. The three remaining Brazilian viruses previously identified as CRF72_BF1 (10BR_MG002, 10BR_MG004, and 10BR_MG008), exhibited mosaic structures highly similar, but not identical, to that of the Spanish viruses and to 10BR_MG003 and 10BR_MG005, with discrepant subtypes in two short genome segments, located in *pol* and gp120^env^. Based on these results, we propose that the 5 viruses from Brazil previously identified as CRF72_BF1 actually belong to two closely related CRFs, one comprising 10BR_MG002, 10BR_MG004, and 10BR_MG008, which keep their CRF72_BF1 designation, and the other, designated CRF122_BF1, comprising 10BR_MG003, 10BR_MG005 and the viruses of the identified Spanish cluster. Three other BF1 recombinant genomes, 2 from Brazil and 1 from Italy, previously identified as unique recombinant forms, were classified as CRF72_BF1. CRF122_BF1, but not CRF72_BF1, was associated with protease L89M substitution, reported to contribute to antiretroviral drug resistance. Phylodynamic analyses estimate the emergence of CRF122_BF1 in Brazil around 1987. Given their close phylogenetic relationship and similar structures, the grouping of CRF72_BF1 and CRF122_BF1 in a CRF family is proposed.

## 1. Introduction

HIV-1 is characterized by high genetic diversity and rapid evolution, derived from elevated mutation and recombination rates. Through these mechanisms, the HIV-1 group M, the causative agent of the AIDS pandemic, has evolved into numerous circulating genetic forms, known as subtypes, of which 10 have been identified (A-D, F-H, K, L), subsubtypes (A1-A6, F1, F2), and circulating recombinant forms (CRFs), 118 of which are currently listed in the Los Alamos HIV Sequence Database (HIV Sequence Database, 2021). Additionally, geographic variants and clusters, some representing substantial proportions of viruses in certain areas, have been identified through phylogenetic analyses within subtypes, subsubtypes, and CRFs (Thomson and Nájera, 2005; Delgado et al., 2015; Delgado et al., 2019). Genetic characterization of HIV-1 variants is of public health relevance, as it allows to track their geographic spread and to estimate their population growth and the efficacy of preventive interventions (Paraskevis et al., 2016; Rife et al., 2017; Vasylyeva et al., 2020). It has also biological and clinical relevance, as different biological properties have been associated with some HIV-1 variants (Kiwanuka et al. 2008; Pérez-Álvarez et al., 2014; Kourí et al., 2015; Venner et al., 2016; Cid-Silva et al., 2018; Song et al., 2019).

The number of CRFs is increasing incessantly, due to both the continuous generation of recombinant forms where diverse HIV-1 variants meet in the same population (Nájera et al., 2002), some of which become circulating through introduction into transmission networks, and the identification of old previously undocumented CRFs. The proportion of CRFs in the HIV-1 pandemic has increased over time, representing around 17% infections in 2010-2015 (Hemelaar et al., 2020). Among CRFs, those derived from subtype B and subsubtype F1 are among the most numerous, 18 of which have been reported in the literature, most of them originated in South America. The most widely circulating CRF from South America is CRF12_BF, which circulates at high prevalences in Argentina and Uruguay, where URFs related to CRF12_BF are frequently found (Thomson et al., 2000; Thomson et al., 2002; Carr et al., 2001). Four other CRF_BFs (numbers 17, 38, 44, and 89) related to CRF12_BF, as evidenced by shared breakpoints and phylogenetic clustering, were subsequently identified in different South American countries (Aulicino et al., 2012; Ruchansky et al., 2009; Delgado et al., 2010; Delgado et al., 2021). Due to their common ancestry and similar structures, these 5 CRFs and related URFs have been proposed to constitute a ‘family’ of recombinant viruses (Thomson and Nájera, 2005; Zhang et al., 2010; Delgado et al., 2021). By contrast, Brazilian CRF_BFs (De Sá Filho et al., 2006; Guimaraes et al., 2008; Sanabani et al., 2010; Pessoa et al., 2014; Reis et al., 2017; Reis et al., 2019) and CRF66_BF (the latter found mainly in Paraguay and Paraguayans living in Spain) (Bacqué et al., 2021) are unrelated to CRF12_BF. Similarly to the viruses of the CRF12_BF family, close relations have been reported between some Brazilian CRF_BFs: CRF28_BF and CRF29_BF (De Sá Filho et al., 2008); and CRF70_BF and CRF71_BF (Pessôa et al., 2014a).

Here we report the spread of a BF1 cluster in Spain whose viruses exhibit a mosaic structure identical to two Brazilian viruses previously identified as CRF72_BF1 (Pessôa et al., 2014b; Pessôa et al., 2016), which would represent a new CRF, with the three other viruses classified as CRF72_BF1 showing highly similar, but not identical, structures. We propose that viruses previously identified as CRF72_BF1 actually belong to two closely related CRFs that constitute a CRF family.

## 2. Materials and methods

### 2.1. Samples

Plasma and whole blood samples from HIV-1-infected individuals were collected in Spain for antiretroviral drug resistance tests and for a molecular epidemiological study. The study was approved by the Committee of Research Ethics of Instituto de Salud Carlos III, Majadahonda, Madrid, Spain. It did not require written informed consent by the study participants, as it used samples and data collected as part of routine clinical practice and patients’ data were anonymized without retaining data allowing individual identification.

### 2.2. PCR amplification and sequencing

An ∼1.4 kb pol fragment in protease-reverse transcriptase (Pr-RT) was amplified from plasma-extracted RNA or from whole blood-extracted DNA by (RT-)PCR followed by nested PCR, as described previously (Delgado et al., 2015) and sequenced with the Sanger method using a capillary automated sequencer. Near full-length genome (NFLG) sequences were obtained for selected samples by RT-PCR/nested PCR amplification from plasma RNA in five overlapping segments and sequenced by the Sanger method, as described (Delgado et al., 2002; Sierra et al., 2005; Cañada et al., 2021). Newly derived sequences are deposited in GenBank under accessions OL982311-OL982317 and OL982320-OL982323.

### 2.3. Phylogenetic sequence analyses

Sequences were aligned with MAFFT v7 (Katoh and Standley, 2013). Initial phylogenetic trees with all Pr-RT sequences obtained by us were constructed via approximate maximum likelihood with FastTree v2.1.10 (Price et al., 2010), using the general time reversible evolutionary model with CAT approximation to account for among-site rate heterogeneity, with assessment of node support with Shimodaira-Hasegawa (SH)-like local support values (Guindon et al., 2010). Subsequent maximum likelihood (ML) trees with sequences of interest were constructed with W-IQ-Tree (Trifinopoulos et al., 2016), using the best-fit substitution model selected by ModelFinder program (Kalyaanamoorthy et al., 2017), with assessment of node support with the ultrafast bootstrap approximation approach (Hoang et al., 2018). Trees were visualized with MEGA v7.0 (Kumar et al., 2016).

Mosaic structures were analyzed by bootscanning (Salminen et al., 1995) with SimPlot v1.3.5 (Lole et al., 1999). In these analyses, trees were constructed using the neighbor-joining method with the Kimura 2-parameter model and a window width of 250 nucleotides. The subtype affiliations of recombinant segments identified with SimPlot, whose breakpoints were more precisely located in the midpoint of transitions between B-F1 subtype-discriminating nucleotides (here defined as those differing between the 75% consensus sequences of subtype B and the Brazilian F1 strain), were further phylogenetically analyzed via ML and Bayesian inference.

These analyses were performed with IQ-Tree; PhyML (Guindon et al., 2010), using the best-fit evolutionary model selected by the SMS program (Lefort et al., 2017) and node support assessment with the approximate likelihood ratio test, SH-like procedure; and MrBayes v3.2 (Ronquist et al., 2012), using the GTR + G + I substitution model, with two simultaneous independent runs and 8 chains 2–5 million generations long, ensuring that both runs reached convergence, as determined by an average standard deviation of split frequencies < 0.01, discarding the first 50% of the trees in the posterior distribution as burn-in. For these analyses, we used as outgroup a reconstructed B-F1 ancestral sequence, considering the phylogenetic relationship between B and F subtypes (Zhu et al., 1998), obtained with IQ-Tree. The use of a reconstructed ancestral sequence as outgroup is similar to the approach used in other studies (Thomson and Fernández-García, 2011; Travers et al., 2004; Seager et al., 2014), to prevent the long branch attraction artifact (Bergsten, 2005) that could be caused by an outgroup whose distance to the ingroup is relatively long compared to the within-ingroup distances. This artifact can result in collapse or substantial decrease in node support of the clades of the ingroup, particularly in short genome segments.

### 2.4. Antiretroviral drug resistance determination

Antiretroviral (ARV) drug resistance was analyzed with the HIVdb program of the Stanford University’s HIV Drug Resistance Database (Rhee et al., 2003; Shafer, 2006).

### 2.5. Temporal and geographical estimations of clade ancestors

The time and the most probable location of the most recent common ancestor (MRCA) of the newly defined CRF were estimated using Pr-RT sequences with the Bayesian Markov chain Monte Carlo (MCMC) coalescent method implemented in BEAST v1.10.4 (Suchard et al., 2018). Before the BEAST analysis, the existence of temporal signal in the dataset was assessed with TempEst v1.5.3 (Rambaut et al., 2016). The BEAST analysis was performed using the SRD06 codon-based evolutionary model (where the 3rd codon position is in a partition different from the 1st and 2nd positions) (Shapiro et al., 2006). We also specified an uncorrelated lognormal relaxed clock and a Bayesian SkyGrid coalescent tree prior (Gill et al., 2013). The MCMCs were run for 20 million generations. The runs were performed in duplicate, and the posterior tree files were combined with LogCombiner v1.10.4. Proper mixing of the chains was assessed with Tracer v1.6, ensuring that effective sample size values of all parameters were >200. The posterior distribution of trees was summarized in a maximum clade credibility (MCC) tree with TreeAnnotator v1.10.4, after removal a 10% burn-in. MCC trees were visualized with FigTree v1.4.2 (Rambaut, http://tree.bio.ed.ac.uk/software/figtree/). Parameter uncertainty was summarized in 95% highest posterior density (HPD) intervals.

## 3. Results

### 3.1. Identification of a HIV-1 cluster of F1 subsubtype in Pr-RT propagating in Spain

In a molecular epidemiological study in Spain, based on Pr-RT sequences, we detected frequent grouping in clusters, several of which were of F1 subsubtype in Pr-RT (Delgado et al., 2015; Delgado et al., 2019). One of them, designated F1_2, which is the focus of the present study, comprised 14 individuals, 13 of them from the region of Galicia, Northwest Spain (Table 1). Years of HIV-1 diagnoses were between 2007 and 2019 and transmission was predominantly heterosexual. Most individuals were Spanish but 3 were Brazilian and 1 was Ukrainian. To determine whether other sequences from databases belonged to this cluster, we performed BLAST searches in the HIV Sequence Database (Los Alamos National Laboratory, 2021), incorporating the most similar sequences in the phylogenetic analyses. This allowed to identify 3 additional sequences that belonged to the F1_2 cluster, from Brazil, Portugal, and Germany respectively (Figure 1). All but two of the viruses collected in Spain and the virus from Germany branched in a subcluster. Viruses from the F1_2 cluster were most closely related to viruses of the Brazilian F1 strain and to Brazilian CRF_BFs with Pr-RT derived from it.

**Table 1.** Epidemiological data of patients and GenBank accessions of sequences.

| Sample ID | City of sample collection | Region of sample collection | Country of origin | Year of HIV diagnosis | Year of sample collection | Gender | Transmission route* | PR-RT GenBank accession | NFLG GenBank accession |
| --- | --- | --- | --- | --- | --- | --- | --- | --- | --- |
| X2592 | Vigo | Galicia | Spain | 2008 | 2008 | F | HT | GU326146 | - |
| X2632 | Ferrol | Galicia | Spain | 2009 | 2009 | M | MSM | GU326158 | KC113006<br>JX140660 |
| X2657 | Vigo | Galicia | Spain | 2009 | 2009 | F | HT | GU326163 | - |
| GA076319 | Vigo | Galicia | Ukraine | 2010 | 2010 | F | HT | OL982311 | - |
| GA099170 | Vigo | Galicia | Switzerland | 2011 | 2011 | F | HT | - | OL982312 |
| GA330265 | Vigo | Galicia | Brazil | 2007 | 2007 | Trans | Sexual | OL982313 | - |
| GA486085 | Ferrol | Galicia | Spain | 2018 | 2018 | F | n.a. | - | OL982314 |
| GA501952 | Vigo | Galicia | Spain | 2014 | 2014 | F | HT | OL982315 | - |
| GA513250 | Vigo | Galicia | Spain | 2012 | 2012 | M | Sexual | OL982316 | - |
| GA522821 | Lugo | Galicia | Brazil | 2019 | 2019 | M | MSM | OL982317 | - |
| GA817166 | Vigo | Galicia | Spain | 2008 | 2016 | F | HT | OL982320 | - |
| GA874035 | Vigo | Galicia | Spain | 2012 | 2012 | M | HT | - | OL982321 |
| GA903064 | Vigo | Galicia | Spain | 2012 | 2012 | F | HT | OL982322 | - |
| NA584314 | Pamplona | Navarre | Brazil | 2017 | 2017 | M | MSM | OL982323 | - |

\*n.a.: datum not available; Trans: transgender; HT: heterosexual; MSM: man who has sex with men; sexual: unspecified sexual.

**Figure 1.**
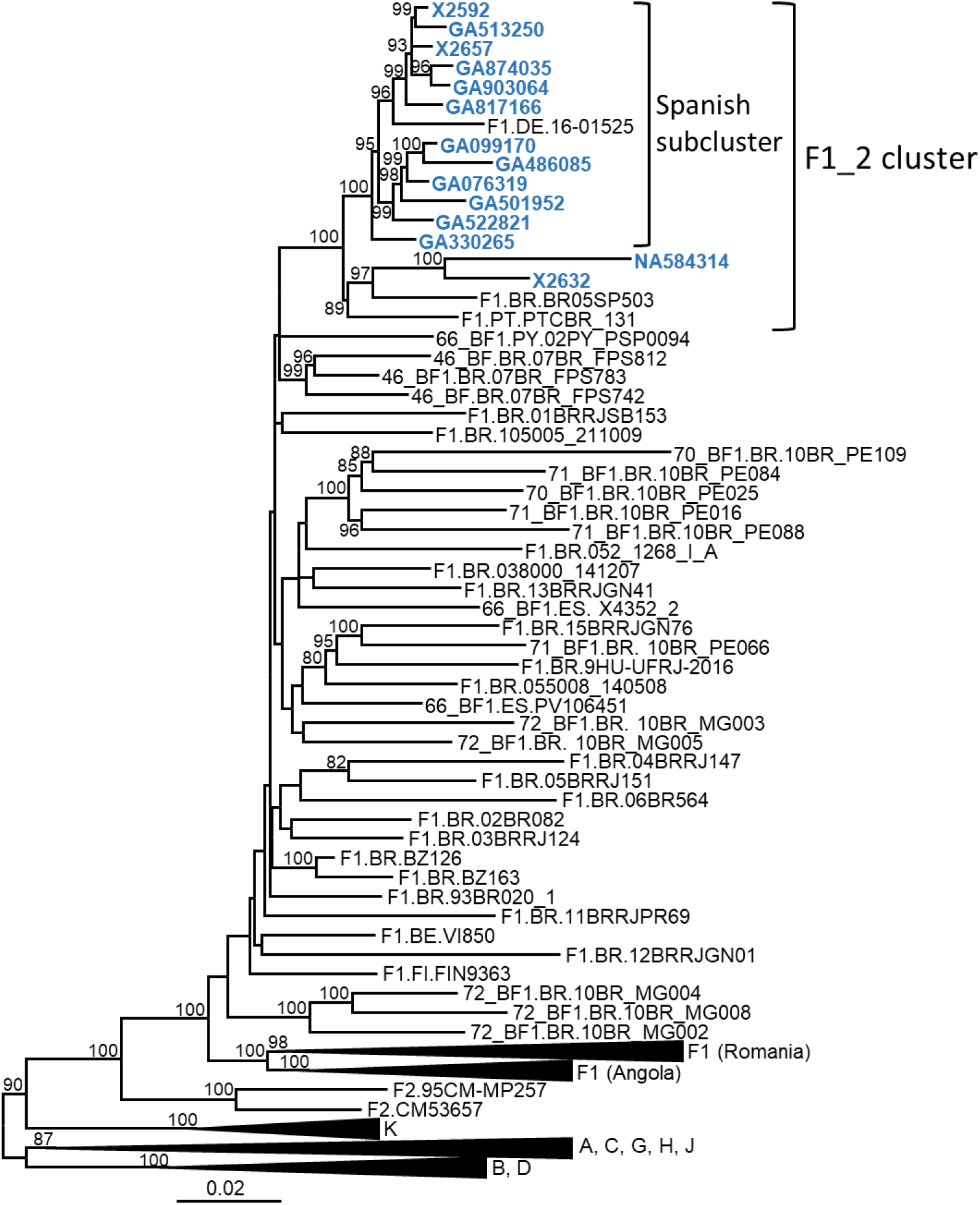
Maximum likelihood phylogenetic tree of Pr-RT sequences of the F1_2 cluster. Names of sequences obtained by us, all collected in Spain, are in blue. Only ultrafast bootstrap values ≥80% are shown. In database sequences, the country of sample collection is indicated before the virus name with the 2-letter ISO country code. BE: Belgium; BR: Brazil; DE: Germany; ES: Spain; FI: Finland; PT: Portugal; PY: Paraguay.

### 3.2. Analyses of NFLG sequences

In order to determine whether the F1_2 cluster was of uniform subtype or recombinant, we obtained NFLG sequences from 3 individuals from 2 cities through amplification from plasma RNA. A fourth NFLG sequence had been obtained previously from virus culture supernatant (Sanchez et al., 2014). Preliminary analyses of the NFLG with Recombination Identification Program (https://www.hiv.lanl.gov/content/sequence/RIP/RIP.html) indicated that the genomes were BF1 recombinant. To determine whether they belonged to a known CRF, we constructed a phylogenetic tree in which genomes of all CRF_BFs were included. The tree showed that viruses of the F1_2 cluster grouped in a strongly supported clade with viruses classified as CRF72_BF1, with two of them, 10BR_MG003 and 10BR_MG005, being the most closely related to the viruses of the F1_2 cluster, and the 3 others, 10BR_MG002, 10BR_MG004, and 10BR_MG008, branching in a sister clade (Figure 2).

**Figure 2.**
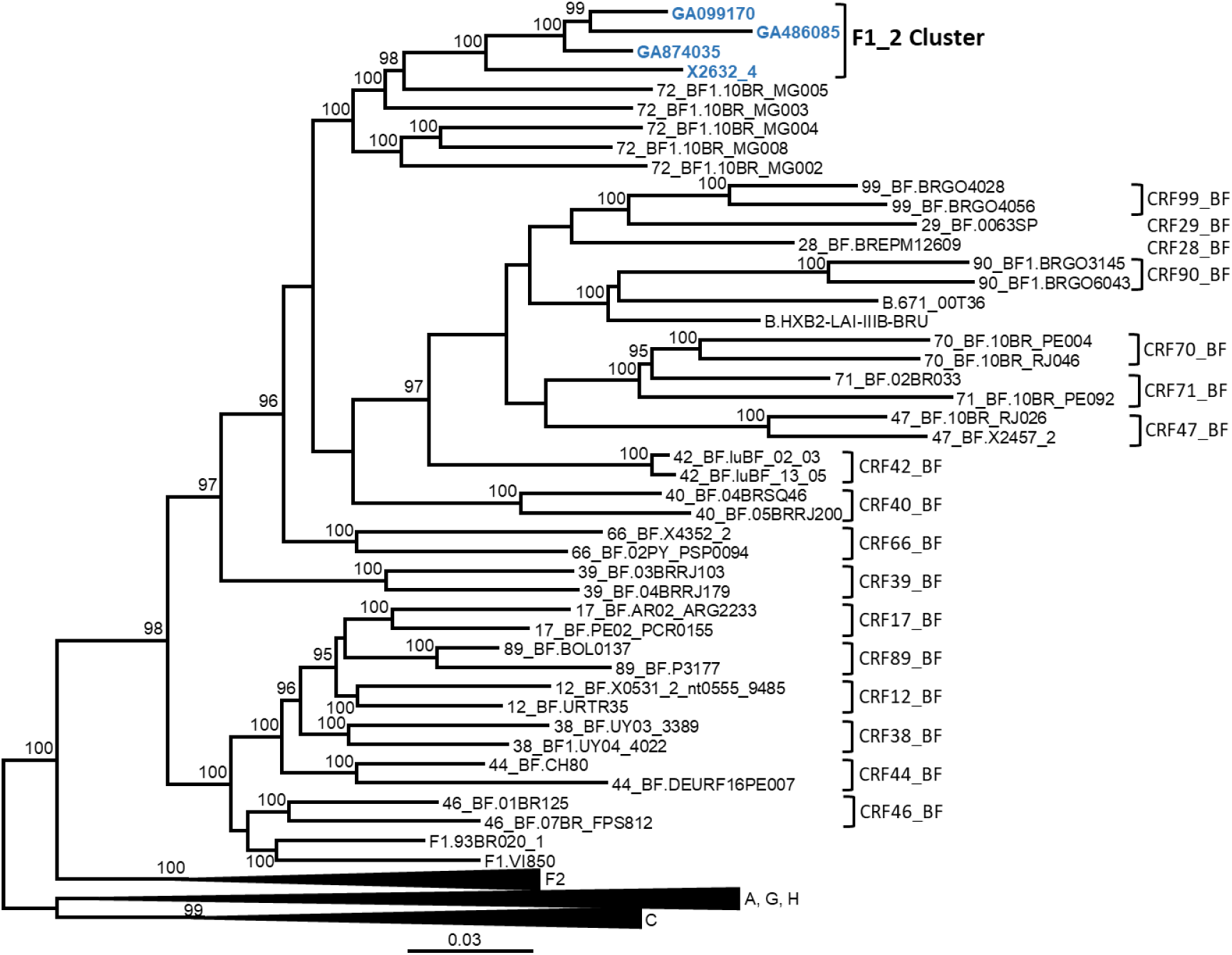
Maximum likelihood tree of NFLG sequences of viruses of the F1_2 cluster. In References of published CRF_BFs and of HIV-1 subtypes are also included in the analysis. Names of sequences obtained by us are in blue. In reference sequences, the subtype or CRF, as classified in the Los Alamos HIV Sequence Database, is indicated before the virus name. Only ultrafast bootstrap values ≥90% are shown.

Bootscan analyses of NFLG sequences showed that the viruses of the F1_2 cluster were BF1 recombinant, exhibiting mosaic structures fully coincident with those of 10BR_MG003 and 10BR_MG005, and slightly different from the three other viruses classified as CRF72_BF1 (Figure 3 and supplementary Figure 1). The differences with these three viruses were observed in a short *pol* segment, around protease-reverse transcriptase (RT) junction, where grouping with subtype references was discrepant, and in the 5’ segment of gp120, where the location of a BF1 breakpoint differed. The mosaic structures determined with bootscanning were confirmed by ML and Bayesian phylogenetic analyses of partial genome segments, which confirmed the coincidence of the mosaic structures of the 4 F1_2 viruses and the Brazilian 10BR_MG003 and 10BR_MG005 viruses and the subtype discrepancy in two genome segments (HXB2 positions 2429-2618 and 6432-6519) of these viruses with 10BR_MG002, 10BR_MG004, and 10BR_MG008 (Figure 4). These analyses, therefore, allowed to determine that viruses of the identified Spanish BF1 cluster, together with the Brazilian viruses 10BR_MG003 and 10BR_MG005, previously classified as CRF72_BF1, belong to a CRF, which was designated CRF122_BF1, which is closely related to, but different from, the three other viruses previously classified as CRF72_BF1, 10BR_MG002, 10BR_MG004, and 10BR_MG008, whose original CRF designation is maintained. The mosaic structures of both CRFs, as inferred from bootscan analyses, ML and Bayesian phylogenetic trees of partial sequences, and examination of intersubtype transitions of subtype-discriminating nucleotides, are shown in Figure 5.

**Figure 3.**
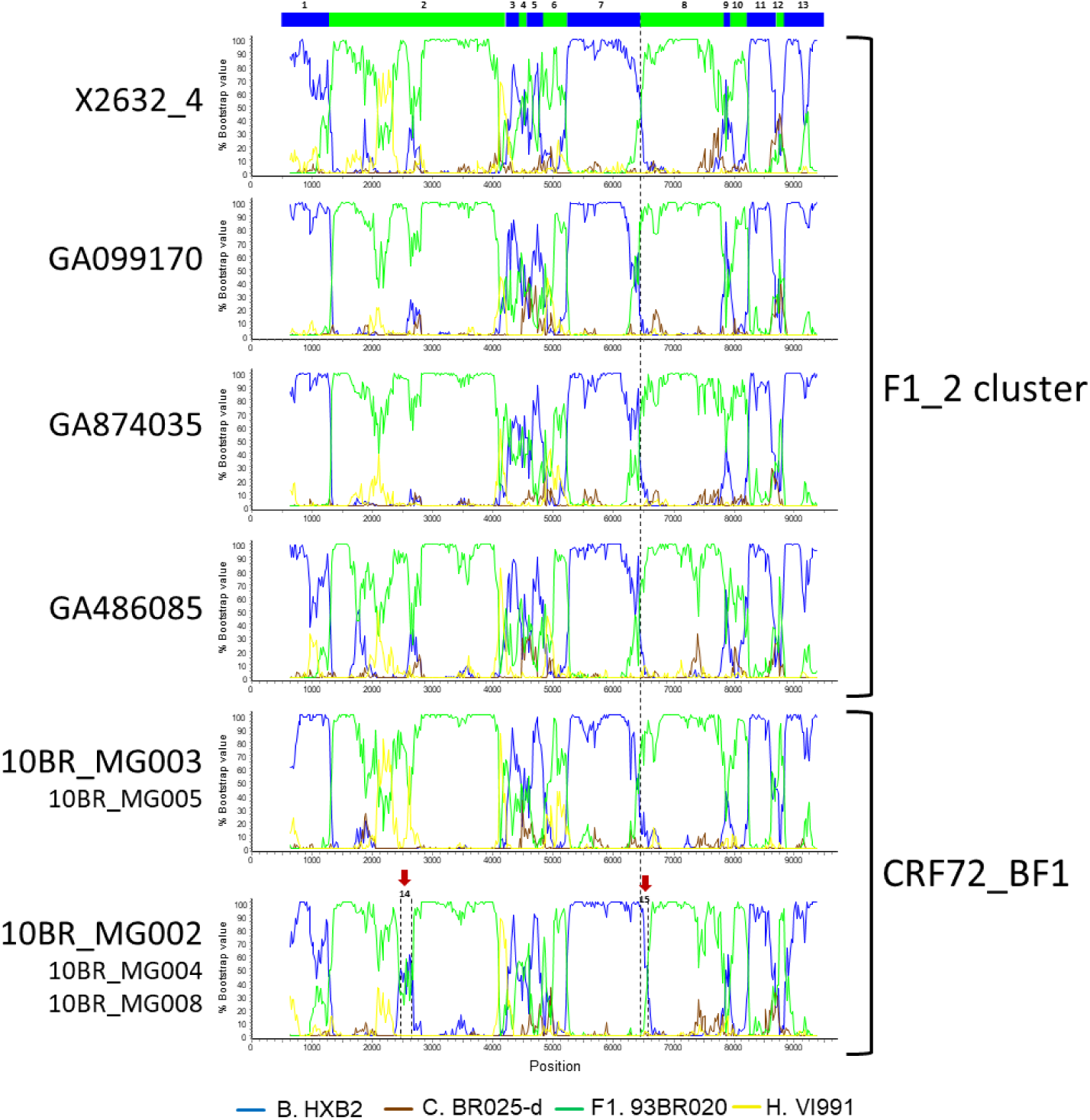
Bootscan analyses of NFLG sequences of viruses of the F1_2 cluster compared to those of viruses previously classified as CRF72_BF1. Bootscan plots of all four F1_2 viruses are shown, together with those of 10BR_MG003 and 10BR_MG002. The bootscan plot of 10BR_MG005 is almost identical to that of 10BR_MG003 and those of 10BR_MG004 and 10BR_MG008 are almost identical to that of 10BR_MG002, and are shown in Supplementary Figure 1. The horizontal axis represents the position in the HXB2 genome of the midpoint of a 250 nt window moving in 20 nt increments and the vertical axis represents bootstrap values supporting clustering with subtype reference sequences. The vertical dashed lines indicate B-F1 breakpoints. The bar on top indicates the numbers of the segments that were further analyzed with ML and Bayesian trees (Figure 4). Two genome segments that appear to group with different subtypes in 10BR_MG002, 10BR_MG004, and 10BR_MG008 relative to 10BR_MG003, 10BR_MG005, and the F1_2 viruses, are signaled with arrows above the bootscan plot of 10BR_MG002 and were also analyzed via ML and Bayesian inference (Figure 4).

**Figure 4.**
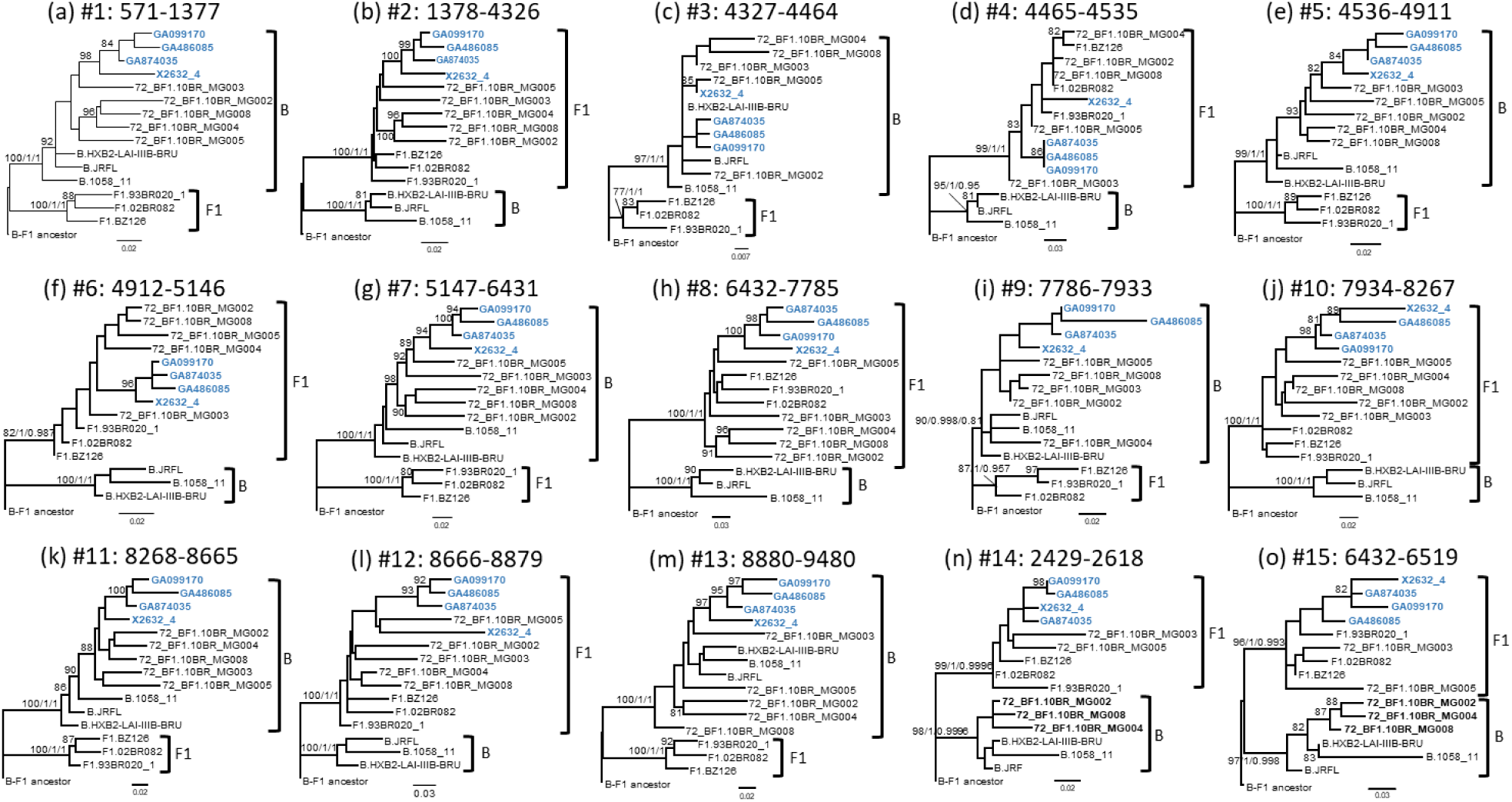
Phylogenetic trees of interbreakpoint genome segments of F1_2 viruses. Breakpoints were defined according to the bootscan analyses and to midpoints of transitions between subtype-discriminating nucleotides, here defined as those where the 75% consensus of subtype B and of the Brazilian variant of subsubtype F1 differ. HXB2 positions delimiting the analyzed segments and their numbers as indicated in Figure 3 are indicated on top of the trees. Sequence names of F1_2 viruses are in blue. Names of subtype reference sequences are preceded by the corresponding subtype name. Sequences of viruses previously classified as CRF72_BF1 were also included. The trees are rooted with a reconstructed B-F1 ancestor. Node supports for B and F1 clades are indicated, in this order, as ultrafast bootstrap value/aLRT SH-like support/posterior probability, which were obtained with IQ-Tree, PhyML, and MrBayes programs, respectively. For the other nodes, only ultrafast bootstrap values ≥80% are indicated. Trees in (n) and (o) (segments 2429-2618 and 6432-6519, respectively) correspond to the segments indicated with arrows in Figure 3 where F1_2 viruses and 10BR_MG003 and 10BR_MG005, on the one hand, and 10BR_MG002, 10BR_MG004, and 10BR_MG008, on the other, appeared to differ in subtype affiliations in the bootscan analyses.

**Figure 5.**
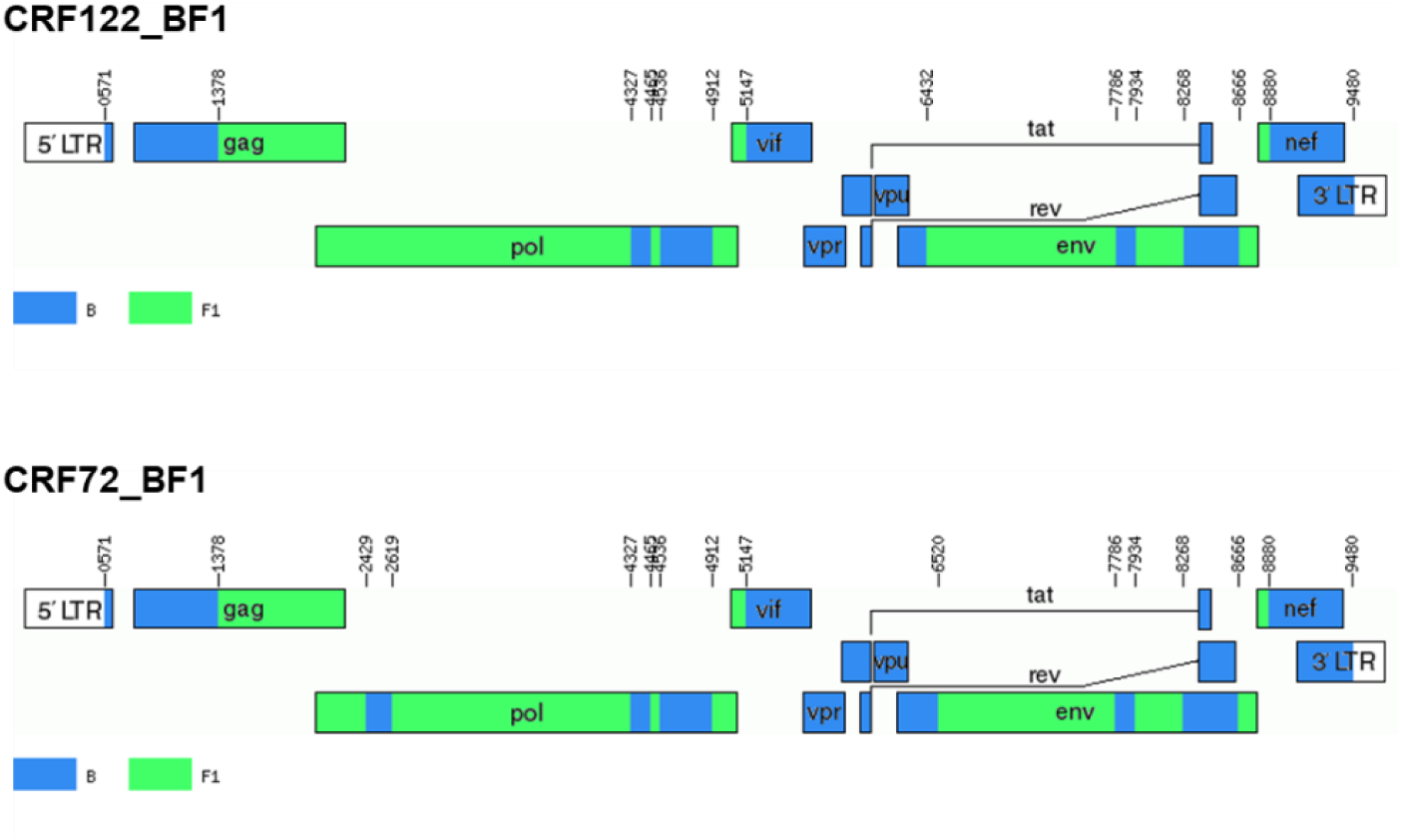
Mosaic structure of CRF72_BF1 and CRF122_BF1. Breakpoint positions are numbered as in the HXB2 genome. The drawing was made using the Recombinant HIV-1 Drawing Tool https://www.hiv.lanl.gov/content/sequence/DRAW_CRF/recom_mapper.html.

Three additional BF1 recombinant NFLGs, originally identified as unique recombinant forms, two from Brazil, 99UFRJ-2 (Thomson et al., 2004) and BREPM1029 (Sa-Filho et al. 2007), and 1 from Italy, IT_BF_PRIN_454 (Bruselles et al., 2009), in their published analyses, exhibited mosaic structures similar to CRF72_BF1 and CRF122_BF1. To determine whether they belonged to one of these CRFs, we constructed a phylogenetic tree with NFLG sequences including the 3 mentioned genomes, which showed that all of them grouped with CRF72_BF1 viruses (Supplementary Figure 2). Bootscan analyses showed mosaic structures of 99UFRJ-2 and IT_BF_PRIN_454 coincident with that of CRF72_BF1; however the bootscan plot of BREPM1029 failed to show clustering with the subtype B reference in the protease-RT junction (HXB2 positions 2429-2618) (Supplementary Figure 3). Examination of subtype-discriminating nucleotides suggested that the 2429-2618 segment was of subtype B in 99UFRJ-2 and BREPM1029, which was confirmed in phylogenetic analyses of this fragment (Supplementary Figure 4a). However, in BREPM1029, the subtype B fragment in the protease-RT junction appeared to be slightly shorter, located between HXB2 positions 2479-2618, which was confirmed in phylogenetic trees (Supplementary Figure 4b). Phylogenetic analyses also showed that in all three genomes the 6432-6519 segment in gp120 was of subtype B, as in CRF72_BF1 (Supplementary Figure 4c). These results allowed to confidently classify 99UFRJ-2 and IT_BF_PRIN_454 as CRF72_BF1 viruses. As to BREPM1029, given its strong phylogenetic clustering with CRF72_BF1 references and its minimal difference in mosaic structure with CRF72_BF1, with a breakpoint displaced only around 50 nt relative to this CRF, it seems reasonable to also classify it as CRF72_BF1, although we cannot definitively discern whether its breakpoint displacement is due to a different recombination event or to mutations occurring near the CRF72_BF1 breakpoint.

### 3.3. Differences in amino acid residues

We analyzed amino acid residues in viral proteins differing between CRF72_BF1 and CRF122_BF1 viruses and conserved within each CRF. We found 10 such amino acid residues (Table 2). One of them is in position 89 of protease, where CRF72_BF1 has leucine, which is the subtype B consensus, while CRF122_BF1 has methionine, which is the F1 subsubtype consensus. Protease L89M substitution has been reported to contribute, together with other protease mutations, to resistance to some protease inhibitor drugs (Calazans et al., 2005; Marcelin et al., 2008; Wensing et al., 2019).

**Table 2.**
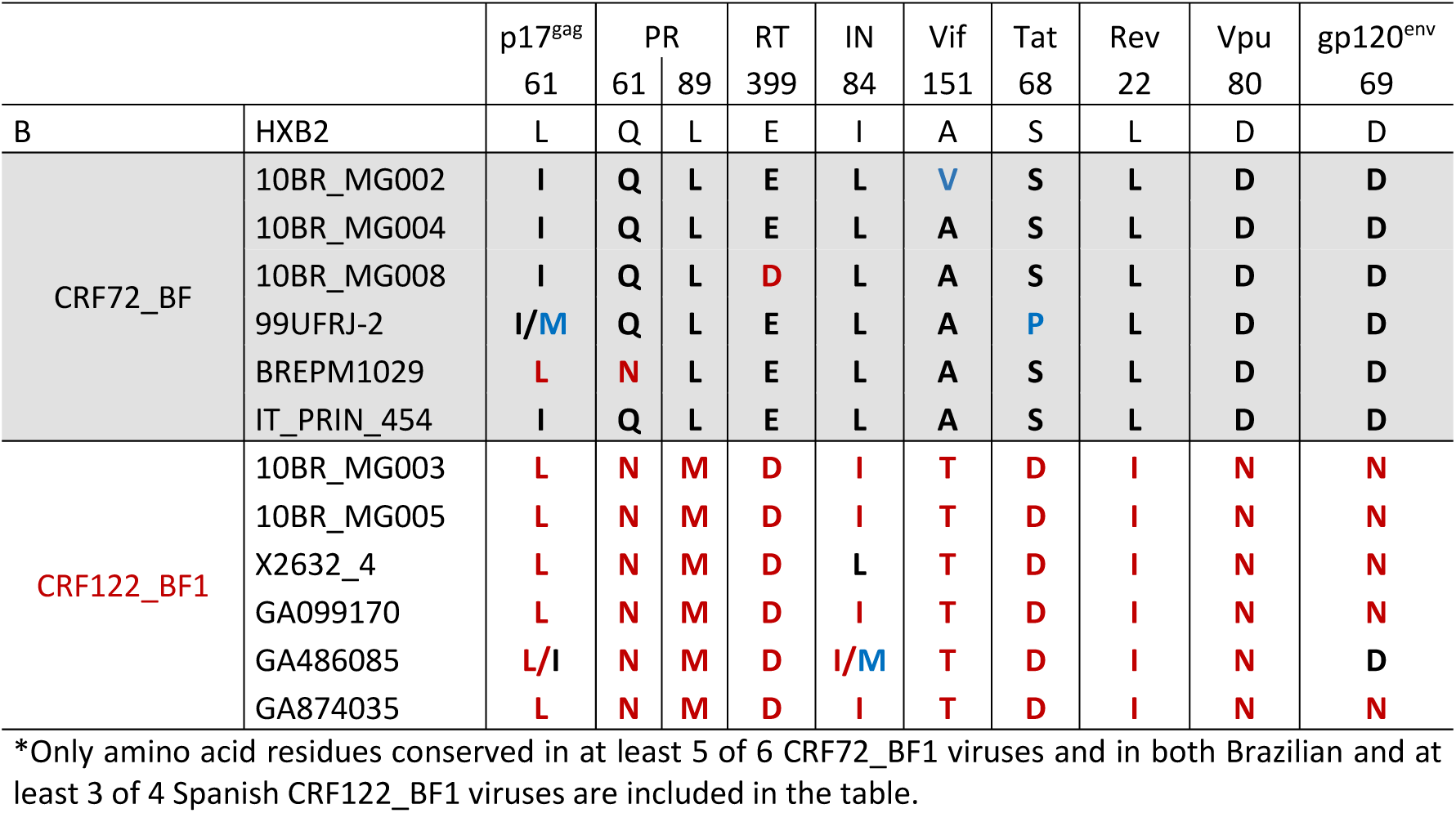
Differences in amino acid residues between CRF72_BF1 and CRF122_BF1*.

### 3.4. ARV drug resistance mutations

Primary ARV drug resistance mutations were detected in 2 CRF122_BF1 viruses, both from Brazil, 1 with L90M protease inhibitor (PI) resistance mutation and the other with D30N PI resistance mutation and M41L, D67N, M184V, and T215Y mutations of resistance to nucleoside RT inhibitors..

### 3.5. Temporal and geographical estimation of CRF122_BF1 origin

To estimate the time and place of origin of CRF122_BF1, Pr-RT sequences where analyzed with the Bayesian coalescent method implemented in BEAST 1.10.4. Prior to this analysis, we performed TempEst analyses to determine whether there was an adequate temporal signal in the dataset. We found that the temporal signal, assessed by the correlation between root-to-tip distance and time, increased by masking the positions of codons with drug resistance mutations in any of the sequences (r^2^ = 0.5265). Therefore, the BEAST analysis was performed with a sequence alignment where these codons had been removed. In this analysis, the substitution rate was estimated at 1.829 × 10^−3^ subs/site/year (95% HPD, 1.118 × 10^−3^ – 2.542 × 10^−3^ subs/site/year) and the time of the MRCA of CRF122_BF1 was estimated around 1987 (95% HPD, 1976-1998), with its most probable location being Brazil (location PP = 0.89) (Figure 6). The introduction of CRF122-BF1 in Spain (according to the MRCA of the Spanish cluster) was estimated in the Galician city of Vigo (location PP=0.992) around 2002 (95% HPD, 1998-2005).

**Figure 6.**
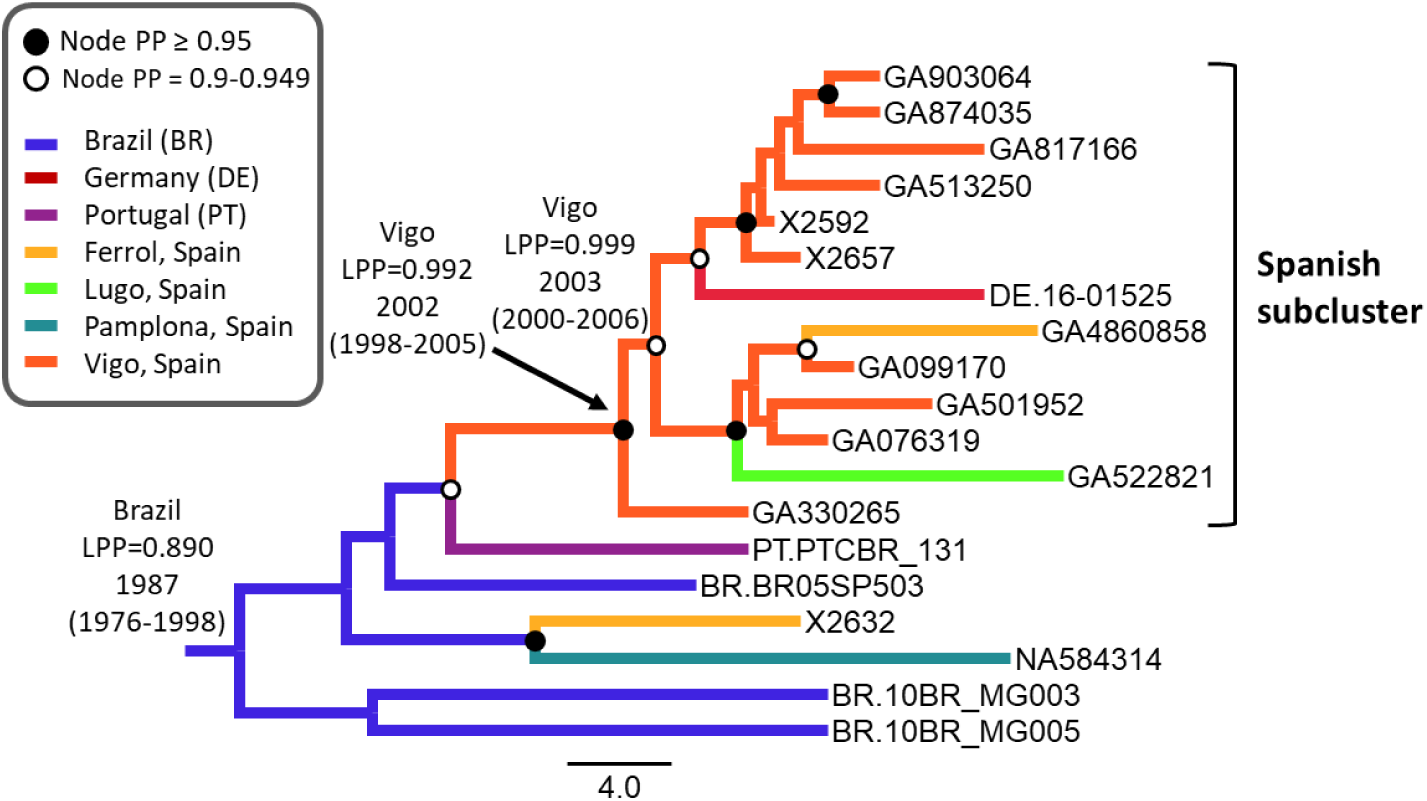
Maximum clade credibility tree of CRF122_BF Pr-RT sequences. Branch colors indicate, for terminal branches, country of sample, and for internal branches, the most probable location country of the subtending node, according to the legend on the upper left. Nodes supported by PP≥0.95 and PP 0.9-0.949 are indicated with filled and unfilled circles, respectively. The most probable locations at the root of the tree and at the node of the Spanish cluster are indicated, together with the PPs supporting each location (LPPs) and the year estimated for the MRCAs (mean value, with 95% HPD interval in parentheses).

## 4. Discussion

The results here presented indicate that the five Brazilian viruses previously classified as CRF72_BF1 actually belong to two closely related CRFs, one of which is circulating in Spain. Consequently, the CRF comprising two Brazilian viruses previously classified as CRF72_BF1 and the four Spanish viruses with coincident mosaic structures are given a new designation, CRF122_BF1, while the three other Brazilian viruses previously classified as CRF72_BF1 keep their original designation. Three other BF1 viruses analyzed in NFLGs originally classified as URFs, two from Brazil and one from Italy, were also classified on the basis of phylogenetic and bootscan analyses as CRF72_BF1. The close relationship between CRF122_BF1 and CRF72_BF1 is one more example of closely related CRFs, with precedents in South America. Other examples are CRF12_BF, CRF17_BF, and CRF89_BF (and more distantly related CRF38_BF and CRF44_BF) (Delgado et al., 2021); CRF28 and CRF29_BF (De Sá Filho et al., 2006); and CRF70_BF and CRF71_BF (Pessôa et al., 2014a).

Failure to realize that the five viruses previously identified as CRF72_BF1 represent two different CRFs may derive from the short segments in which both CRFs differ in subtypes. These differences may be missed if bootscan analyses are performed using window widths much greater than the length of the recombinant fragment. We have also noticed that jpHMM (Schultz et al., 2009), used in a previous study to analyze CRF72_BF1 genomes (Pessôa et al., 2006), often fails to detect short recombinant segments.

Given the close relationship and partial coincidence in mosaic structures of CRF72_BF1 and CRF122_BF1, we propose that they are members of a CRF family, similarly to the CRF family of BF1 recombinant viruses from South America comprising CRFs numbers 12, 17, 38, 44, and 89 (Delgado et al., 2021). The grouping of some closely related HIV-1 recombinants derived from a common recombinant ancestor in families was proposed by us (Thomson and Nájera, 2005) and others (Zhang et al., 2010). Proposed families of CRFs with close phylogenetic relations, shared parental strains, and partially coincident breakpoints are indicated in the phylogenetic tree of Supplementary Figure 5.

The estimated origin of CRF122_BF1 around 1987 is consistent with the estimated origin of the Brazilian F1 strain (around 1977) (Bello et al., 2006) and similar to those of other South American CRF_BFs (CRF12, CRF28/29, CRF38, CRF89, and CRF90) reported in the literature (Bello et al., 2010; Ristic et al., 2011; Reis et al., 2017; Delgado et al, 2021), but younger than some other estimates for CRF12_BF in the 1970s (Dilernia et al., 2011; Delgado et al., 2021) and older than the estimates for CRF99_BF, around 1993 (Reis et al., 2017).

The correct classification of HIV-1 genetic forms is important, since even relatively minor genetic differences in viral genomes may result in important biological differences. Examples in HIV-1 are CXCR4 coreceptor usage in CRF14_BG, which is associated with only four amino acid residues in the Env V3 loop (Pérez-Álvarez et al, 2014), all or most of which are absent in viruses of the closely related CRF73_BG (Fernández-García et al., 2016), which has a very similar mosaic structure; and differences in pathogenic potential or therapeutic response associated with clusters within HIV-1 CRF01_AE (Song et al., 2019) and F1 subsubtype (Cid-Silva et al., 2018). Here we show that CRF122_BF1, but not CRF72_BF1, has the protease L89M substitution, that has been reported to contribute, together with other protease mutations, to resistance to tipranavir/ritonavir (Marcelin et al., 2008) and, within an F1 subsubtype background, to resistance to other protease inhibitor drugs (Calazans et al., 2005).

CRF122_BF1 represents one more example of a CRF of South American ancestry first identified in Western Europe. Others are CRF42_BF (Struck et al., 2015), CRF47_BF (Fernández-García et al., 2010), CRF60_BC (Simonetti et al., 2014), CRF66_BF (Bacqué et al., 2021), and CRF89_BF (Delgado et al., 2021). This may derive from the increasing migratory flows from South America to Europe and from the relatively low number of

HIV-1 sequences available in some South American countries (Bacqué et al., 2021). Therefore, HIV-1 molecular epidemiological studies in Europe may contribute to a better knowledge of the HIV-1 epidemics in South America.

In summary, we show that viruses of a BF1 recombinant cluster of Brazilian ancestry circulating in Spain exhibit a mosaic structure that is fully coincident with that of two Brazilian viruses previously classified as CRF72_BF1, and highly similar, but not identical, to that of three other Brazilian viruses also classified as CRF72_BF1. Therefore, we propose a new CRF designation, CRF122_BF1, for the viruses of the Spanish cluster and the two Brazilian viruses with coincident structures, which together with CRF72_BF1 would constitute a CRF family. The accurate genetic characterization of HIV-1 variants is important to determine their associated biological features and to track their epidemic spread.

## Data Availability

All sequences produced in the present study have been deposited in GenBank, under accessions OL982311-OL982317 and OL982320-OL982323. Epidemiological data are available in the manuscript.

## Acknowledgments

We thank José Antonio Taboada, from Consellería de Sanidade, Xunta de Galicia, for his support of this study, and the personnel at the Genomic Unit, Instituto de Salud Carlos III, for technical assistance in sequencing.

## Author contributions

MT and ED conceived the study and supervised the experimental work. JC-G, ED, and MT processed sequences and performed phylogenetic and phylodynamic analyses. HG performed data curation. JC-G, SB, MS and EG-B performed experimental work. AO, JJC, CM, AM, PO, MJG, and CE obtained samples and epidemiological data from patients. MT, JC-G, ED, and HG wrote the manuscript. All authors read and approved the manuscript.

