## Supplementary figures for "Viruses previously identified in Brazil as belonging to HIV-1 circulating recombinant form (CRF) 72_BF1 represent two closely related CRFs, one of which, designated CRF122_BF1, is also circulating in Spain"

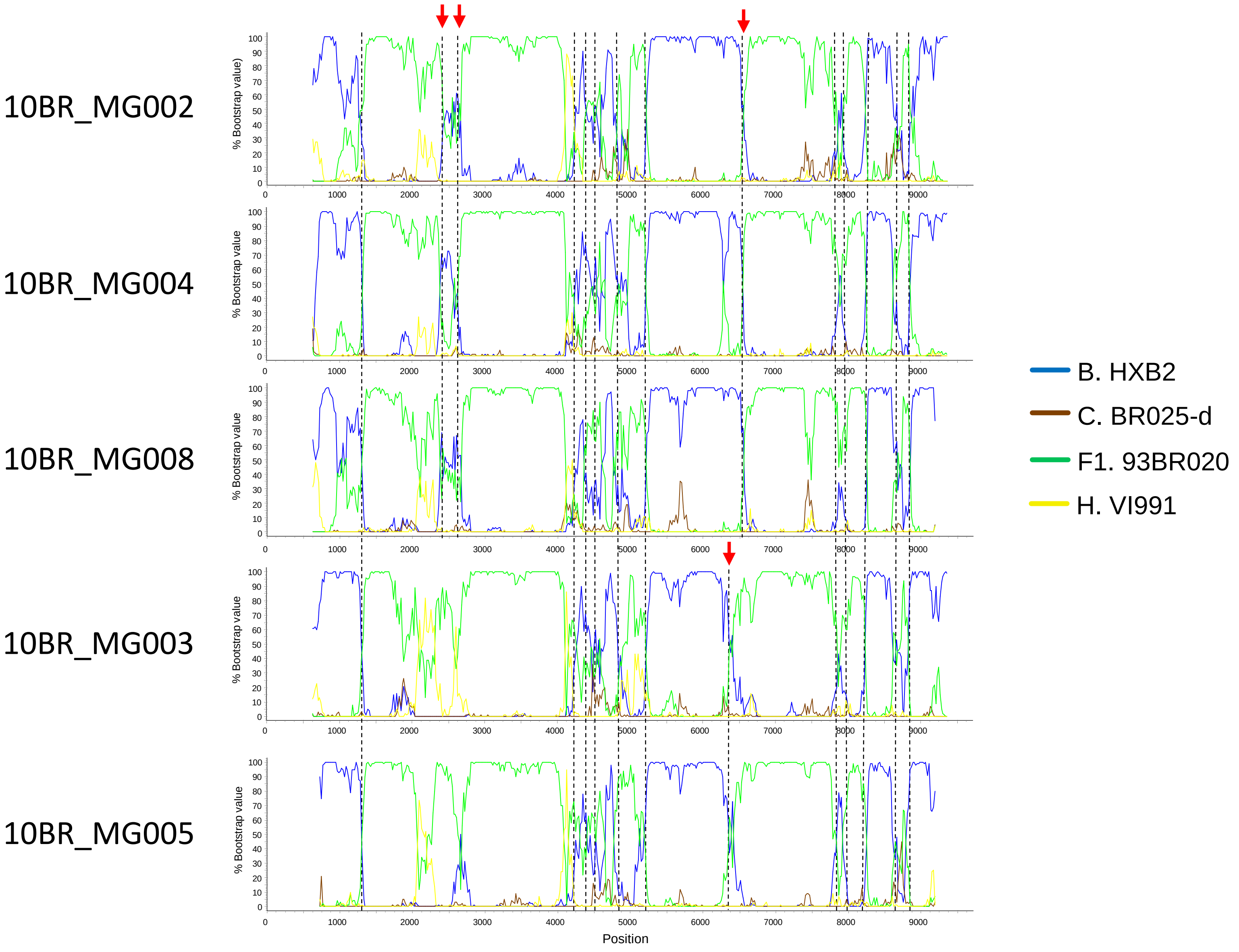


**Supplementary Figure 1. Bootscan analyses of NFLG sequences of 5 viruses previously classified as CRF72_BF1.** The horizontal axis represents the position in the HXB2 genome of the midpoint of a 250 nt window moving in 20 nt increments and the vertical axis represents bootstrap values supporting clustering with subtype reference sequences. Vertical dashed lines indicate breakpoints. Breakpoints differing between 10BR_MG002, 10BR_MG004, and 10BR_MG008, on the one hand, and 10BR_MG003 and 10BR_MG005, on the other, are signaled with arrows.

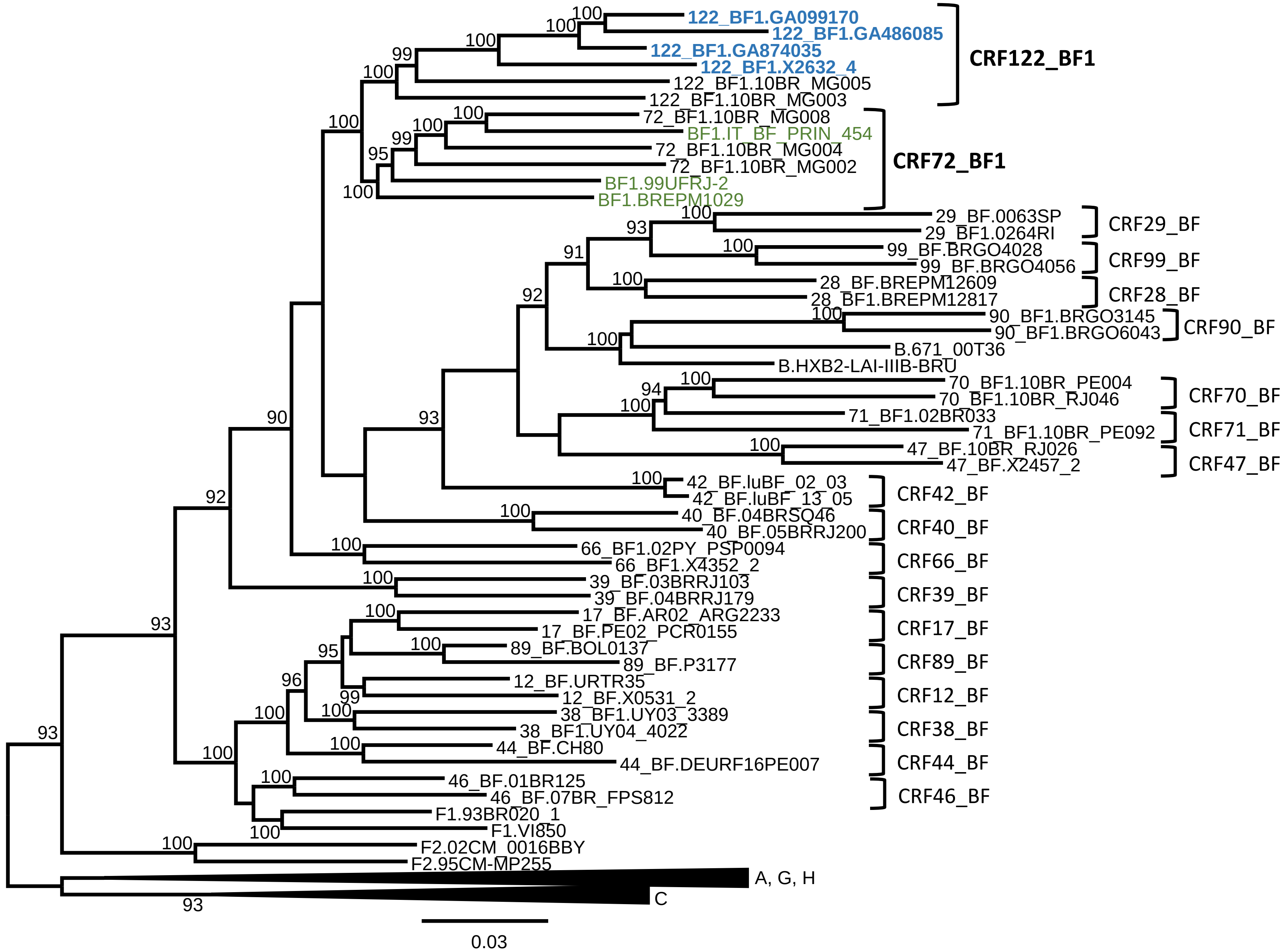


**Supplementary Figure 2. Maximum likelihood tree of NFLG sequences of 99UFRJ-2, BREPM1029, and IT_BF_PRIN_454, with CRF_BF references.** The tree was constructed with IQ-Tree and the node support values are ultrafast bootstrap values. It is observed that 99UFRJ-2, BREPM1029, and IT_BF_PRIN_454 group in a strongly supported clade with CRF72_BF1 viruses.

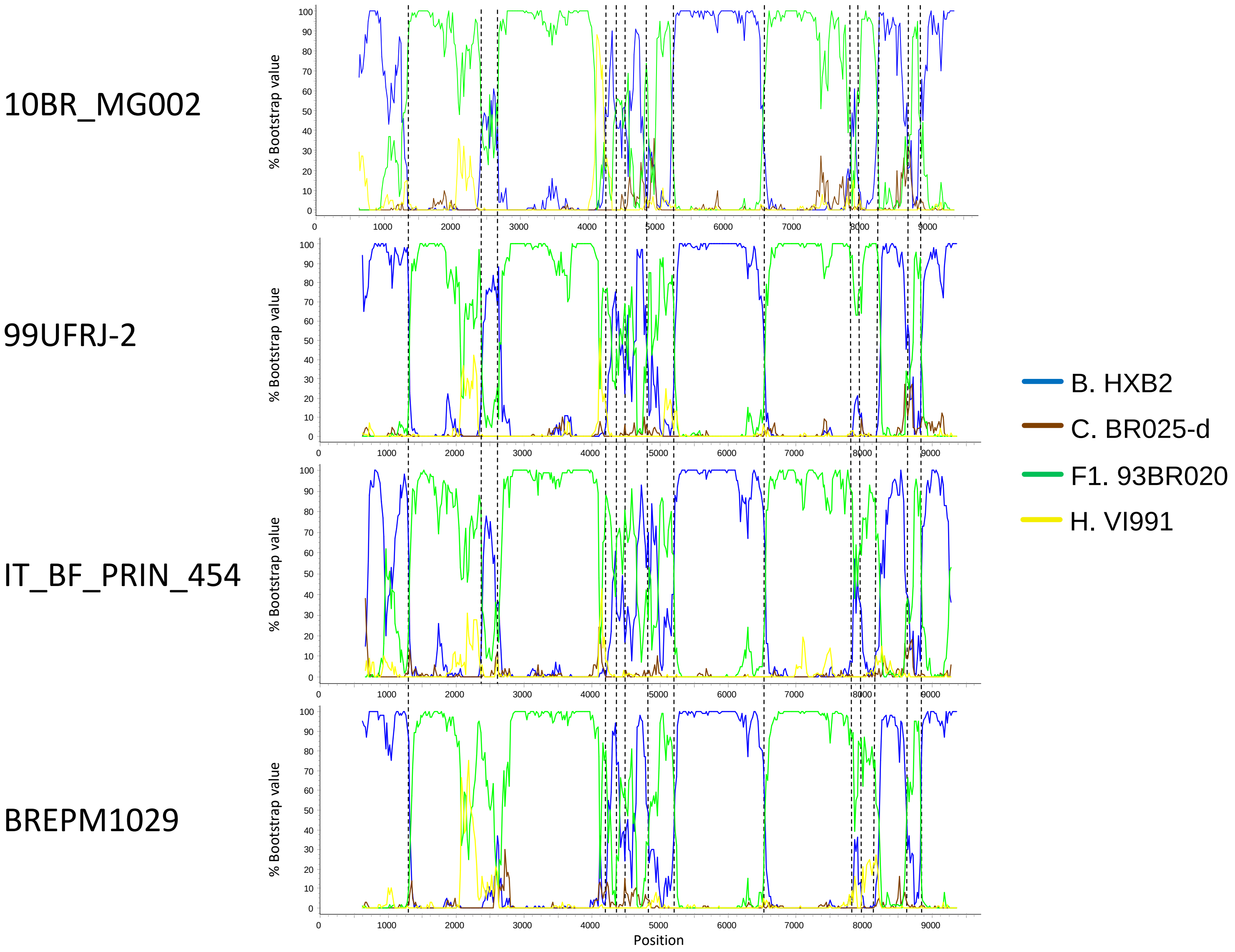


**Supplementary Figure 3. Bootscan analyses of NFLG sequences of 99UFRJ-2, BREPM1029, and IT_BF_PRIN_454.** The bootscan plot of 10BR_MG002 is also shown for comparison. The horizontal axis represents the position in the HXB2 genome of the midpoint of a 250 nt window moving in 20 nt increments and the vertical axis represents bootstrap values supporting clustering with subtype reference sequences. Vertical dashed lines indicate breakpoints

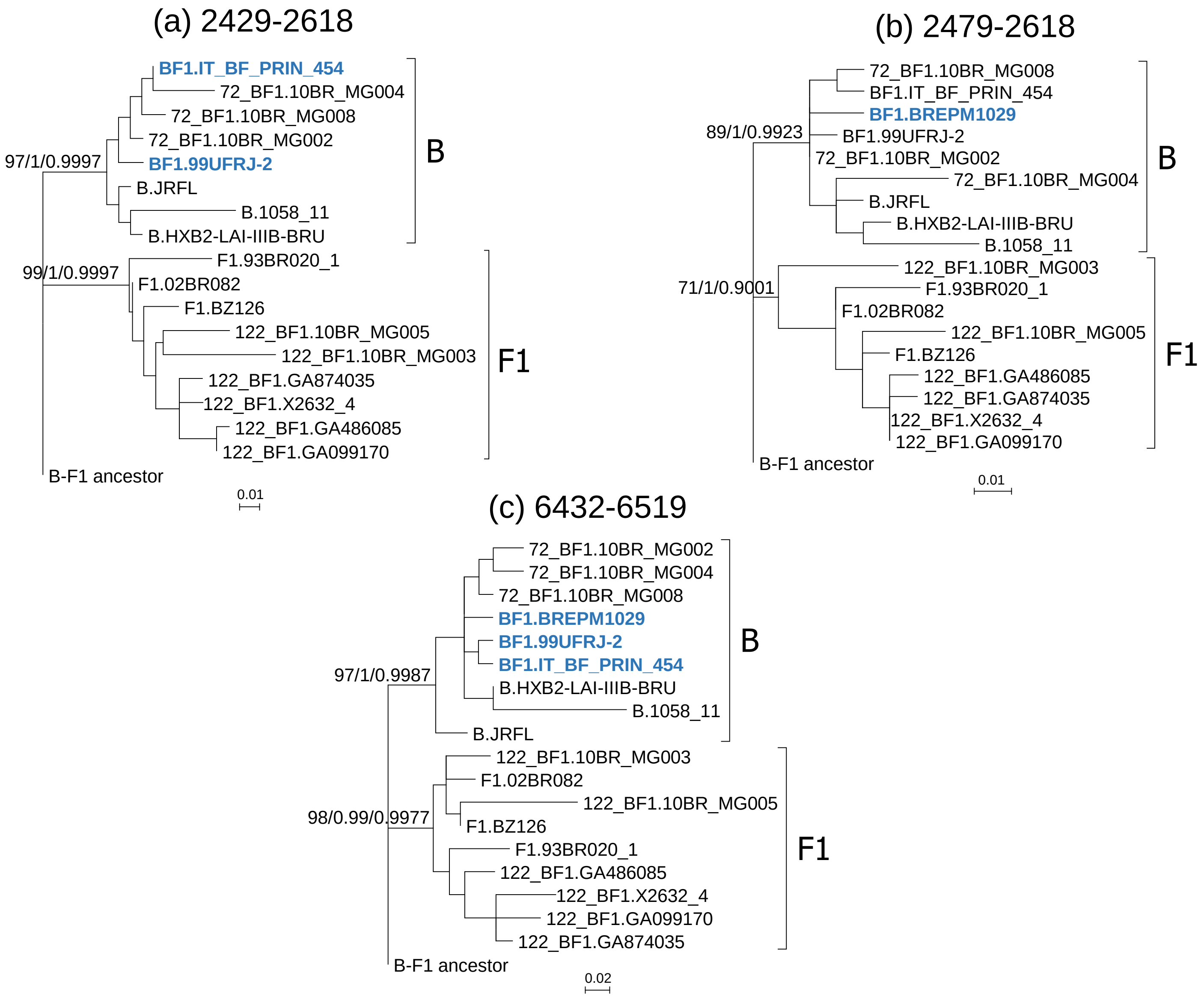


**Supplementary Figure 4. Phylogenetic trees of 99UFRJ-2, BREPM1029, and IT_BF_PRIN_454 in genome segments where CRF72_BF1 and CRF122_BF1 differ in subtype.** HXB2 positions are indicated above the trees. Node supports of B and F1 clades are indicated, in this order, as ultrafast bootstrap value/aLRT SH-like support/posterior probability, obtained with IQ-Tree, PhyML, and MrBayes programs, respectively. In the protease-RT junction segment, given a slightly different breakpoint location, BREPM1029 is analyzed separately.

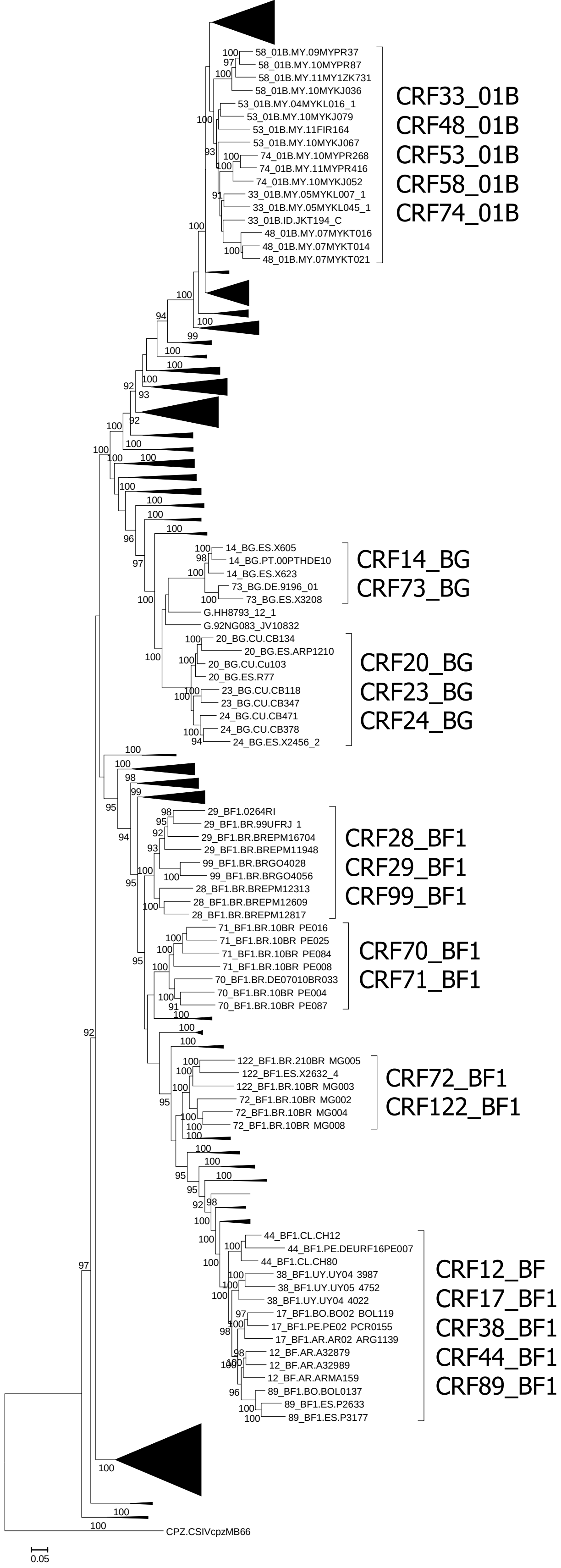


**Supplementary Figure 5. Maximum likelihood tree of NFLG sequences of HIV-1 CRFs.** Ultrafast bootstrap values ≥90% are shown. CRF groups that show strong phylogenetic clustering, derive from the same parental strains, and have partially coincident mosaic structures, are indicated with brackets. These groups are proposed to constitute CRF families.
